# From Christmas sex to winter intimacy: three decades of birth seasonality, sex ratio dynamics, and fertility change in South Africa, 1994–2024

**DOI:** 10.64898/2026.07.12.26357853

**Authors:** Rumbidzai Masukume, Pugie T Chimberengwa, Gwinyai Masukume, Grażyna Liczbińska, Victor Grech, Witness Mapanga

## Abstract

**Background:** Since South Africa’s democratic transition in 1994, the country has undergone profound social, demographic and public health change. We analysed national recorded live-birth data from 1994–2024 to identify major signals of population reproduction.

**Methods:** Monthly recorded live births from January 1994 to December 2024 were obtained from Statistics South Africa. Birth seasonality, sex ratio at birth (SRB) [male/total live births] and annual recorded live births were analysed using time-series and forecasting methods.

**Results:** From 1994–2014, September was the peak birth month in all 21 years, consistent with conceptions during the Christmas–New Year holiday period nine months earlier. From 2015 onwards, March became the most frequent peak month, with April emerging as the peak month in 2024, indicating a shift towards winter conceptions. The SRB declined to 49.996% in June 2021 (95% prediction interval 50.165%–50.749%) and remained below the lower prediction bound from May to July 2021 (combined *p*<0.001). November 2021 recorded the highest monthly SRB in the 31-year study period (50.983%), exceeding the upper 95% prediction interval. Annual recorded live births peaked at 1,112,378 in 2008 and declined to 798,556 in 2024; births from 2022–2024 fell below the 95% confidence interval of the historical trend.

**Conclusions:** Three prominent demographic signals emerged: a shift from Christmas holiday conceptions towards winter conceptions; a rare inversion (SRB <50%) and sustained depression of the SRB during May–July 2021, occurring within the 3–5-month stress-sensitive window after the January 2021 Beta-wave mortality peak, followed by the highest monthly SRB of the study period in November, nine months after the easing of COVID-19 restrictions in February 2021; and an accelerated decline in annual recorded live births after 2021, culminating in the lowest level observed in 2024. These findings indicate changes in reproductive timing, stress-sensitive sex-ratio patterning and fertility in South Africa.

## Introduction

Seasonal variation in human births has been documented for centuries and remains a useful indicator of population-level reproductive behaviour. Given the approximately nine-month interval between conception and live birth (Jukic et al., 2013), temporal patterns in birth occurrence provide indirect evidence of conception timing and the underlying social, environmental, and biological determinants of human reproduction. These conditions include climate, infectious disease, work patterns, holidays, religious observance, food security, migration and changes in sexual behaviour (Cowgill, 1966, Lam and Miron, 1991, Dorélien, 2016, Ellison and Valeggia, 2005, Régnier-Loilier, 2010, Alcaide et al., 2023, Barreca et al., 2018, Grech et al., 2003, Zammit and Grech, 2020). Modernisation has reduced the amplitude of some historical birth cycles, but seasonality has not disappeared. Rather, seasonal patterns often shift as societies change (Dorélien, 2016, Régnier-Loilier, 2010, Alcaide et al., 2023, Barreca et al., 2018).

South Africa is a particularly informative setting in which to examine these processes. Historical analyses showed that South African births peaked in September during much of the twentieth century. Cowgill described this pattern using data from 1935 to 1958, and Lam and Miron later reported a September peak using South African vital registration data from 1950 to 1984 (Cowgill, 1966, Lam and Miron, 1991). Dorélien, in a wider analysis of sub-Saharan Africa, noted the same South African pattern and showed that birth seasonality remains common in African populations (Dorélien, 2016). A September birth peak implies elevated conceptions during December and early January, making the Christmas and New Year period a plausible social anchor for reproductive timing.

Since South Africa’s democratic transition in 1994, the country has undergone profound social, economic, demographic, and public health transformation. Because birth seasonality is sensitive to changes in population behaviour and living conditions, these societal transitions may have altered long-standing patterns of human reproduction. A recent parliamentary review marking three decades of democracy framed this period as one of democratic transformation, institutional rebuilding and continuing social challenge (Parliament of the Republic of South Africa, 2024). Mayosi and Benatar described considerable social progress after apartheid alongside persistent inequality and health-system challenges (Mayosi and Benatar, 2014). Electrification, housing change, migration, urbanisation, antiretroviral therapy scale-up and shifting household conditions altered daily life (Mayosi and Benatar, 2014, Ukoba et al., 2025). These changes may plausibly influence reproductive behaviour and timing, although such effects cannot be directly inferred from aggregate birth data.

In addition to birth seasonality and birth counts, population-level reproductive dynamics may also be reflected in variation in the sex ratio at birth (SRB). The SRB, here defined as male live births as a percentage of total live births, is a particularly sensitive demographic signal. Typically, male live births slightly outnumber female live births, and international reference-level work has estimated national and regional SRB levels (Chao et al., 2019, James, 1987). South African work has also suggested that SRB may act as a sentinel indicator of population health (Grech and Masukume, 2016).

The COVID-19 pandemic, declared by the World Health Organization in March 2020 (Cucinotta and Vanelli, 2020), introduced a distinct and acute shock. South Africa experienced multiple epidemic waves, with particularly severe mortality during the second wave, when the Beta variant predominated (Bradshaw et al., 2022, Tegally et al., 2021, Jassat et al., 2021, Nyagupe et al., 2023). The pandemic also affected maternal and perinatal health, mental health, economic security and the functioning of health services (Fawcus et al., 2024, Hirachund et al., 2024, Dladla-Jaca et al., 2024). Such shocks have been hypothesised in previous literature to be associated with short-term variation in birth outcomes, including SRB, although evidence remains heterogeneous across populations and events (Catalano and Bruckner, 2006, Grech, 2014, Helle et al., 2009). First, acute population stress has been proposed to alter the SRB, potentially via differential fetal loss or survival patterns, a phenomenon discussed in the sex-ratio literature and sometimes interpreted within, though not exclusively explained by, the Trivers–Willard framework (Grech and Masukume, 2016, Trivers and Willard, 1973, Garenne, 2002, Masukume and Grech, 2015, Masukume et al., 2022, Catalano et al., 2006). Second, mortality, uncertainty and economic disruption can alter fertility behaviour, changing the number and timing of births at the population level (Pomar et al., 2022, Sobotka et al., 2024).

A previously published analysis showed that the onset of COVID-19 in South Africa was followed by a June 2020 decline and inversion of the SRB (Masukume et al., 2022). Whether further perturbations occurred after the first wave has not been evaluated using more recent data available through 2024. Taken together, birth seasonality, SRB and annual live-birth counts provide complementary indicators of population reproductive dynamics and responses to social change. This study integrates three related but analytically distinct signals: birth seasonality, SRB and annual recorded live births. The aim was to characterise prominent hallmark changes in these from 1994 to 2024 using descriptive and time-series methods, explicitly treating each outcome as a separate demographic signal rather than assuming a shared causal structure across outcomes.

## Methods

### Data source

Monthly recorded live births from January 1994 through December 2024, the most recent data available, were obtained from Statistics South Africa recorded live birth releases and appendices (Statistics South Africa, 2025). The submitted dataset included monthly counts of male live births, female live births and total live births. Annual totals were derived by summing monthly totals. The analysis used aggregate public-domain data only. Statistical analyses were performed using Stata/SE version 17 (StataCorp LLC, College Station, TX, USA).

### Birth seasonality

Monthly birth counts were converted to average births per day to account for differences in month length. Within each calendar year, months were ranked from highest to lowest according to average daily births, with rank 1 representing the highest average daily birth intensity and rank 12 the lowest.

Estimated conception month ranks were obtained by shifting birth months approximately nine months earlier. This ecological approximation was used to examine population-level seasonal patterns and does not account for variation in gestational duration between individual pregnancies.

Changes in birth seasonality were assessed by comparing monthly birth-rank distributions before (1994–2020) and from 2021 onwards. Particular attention was given to the March, April and September birth peaks, which approximately correspond to conceptions during the June, July and December holiday periods. Differences in the distributions of monthly ranks between the two periods were assessed using the Wilcoxon rank-sum (Mann–Whitney) test.

### Sex ratio at birth

The SRB was calculated as the percentage of male live births among total live births for each month. To estimate the expected SRB during the COVID-19 and post-pandemic period, forecasts were generated using data from January 2015 to December 2019. This five-year period (60 monthly observations) represents the recent pre-pandemic baseline while excluding any influence of the COVID-19 pandemic and is consistent with the duration commonly used in COVID-19 forecasting studies (Achilleos et al., 2022).

Stationarity of the pre-pandemic SRB series was assessed using the Augmented Dickey-Fuller test before model fitting. Autoregressive moving average (ARMA) models with autoregressive and moving-average orders ranging from 0 to 5 were fitted to the pre-pandemic series. Candidate models were compared using Akaike’s Information Criterion (AIC), and the model with the lowest AIC was selected (Supplementary Table 1).

The selected ARMA model was used to generate forecasts of monthly SRBs for January 2020 through December 2024. The 95% prediction intervals were calculated using Stata’s predict command with the mse option (StataCorp, 2019). Months with observed SRBs falling outside the 95% prediction intervals were identified directly from the model forecasts. For the consecutive deviations observed during May–July 2021, two-sided probabilities derived from the forecast distribution were combined using Fisher’s method.

### Annual live births

Annual recorded live births were analysed descriptively using annual counts from 1994 to 2024. To assess whether births during the pandemic and post-pandemic period differed from historical expectations, a parsimonious linear regression model was fitted to pre-pandemic annual births (1994–2019), with calendar year as the independent variable. The fitted model was used to estimate expected annual births for 2020–2024, and 95% confidence intervals around the expected trend were derived from the regression model. Observed annual births were compared with the expected values and corresponding confidence intervals. A simple linear model was chosen because it provides a transparent and reproducible benchmark for contextual comparison rather than a definitive demographic forecasting model.

### Ethics

Ethics approval was not required because the study used only anonymised, aggregated, publicly available data and involved no individual-level or identifiable human participant information.

## Results

### Birth seasonality

From 1994 to 2014, September was the highest-ranked birth month in every year, indicating a highly stable historical pattern of birth seasonality (Fig. 1A). Based on an approximate nine-month gestational interval, this corresponds to conceptions occurring predominantly during December and early January (Fig. 1B).

**Figure 1A.**
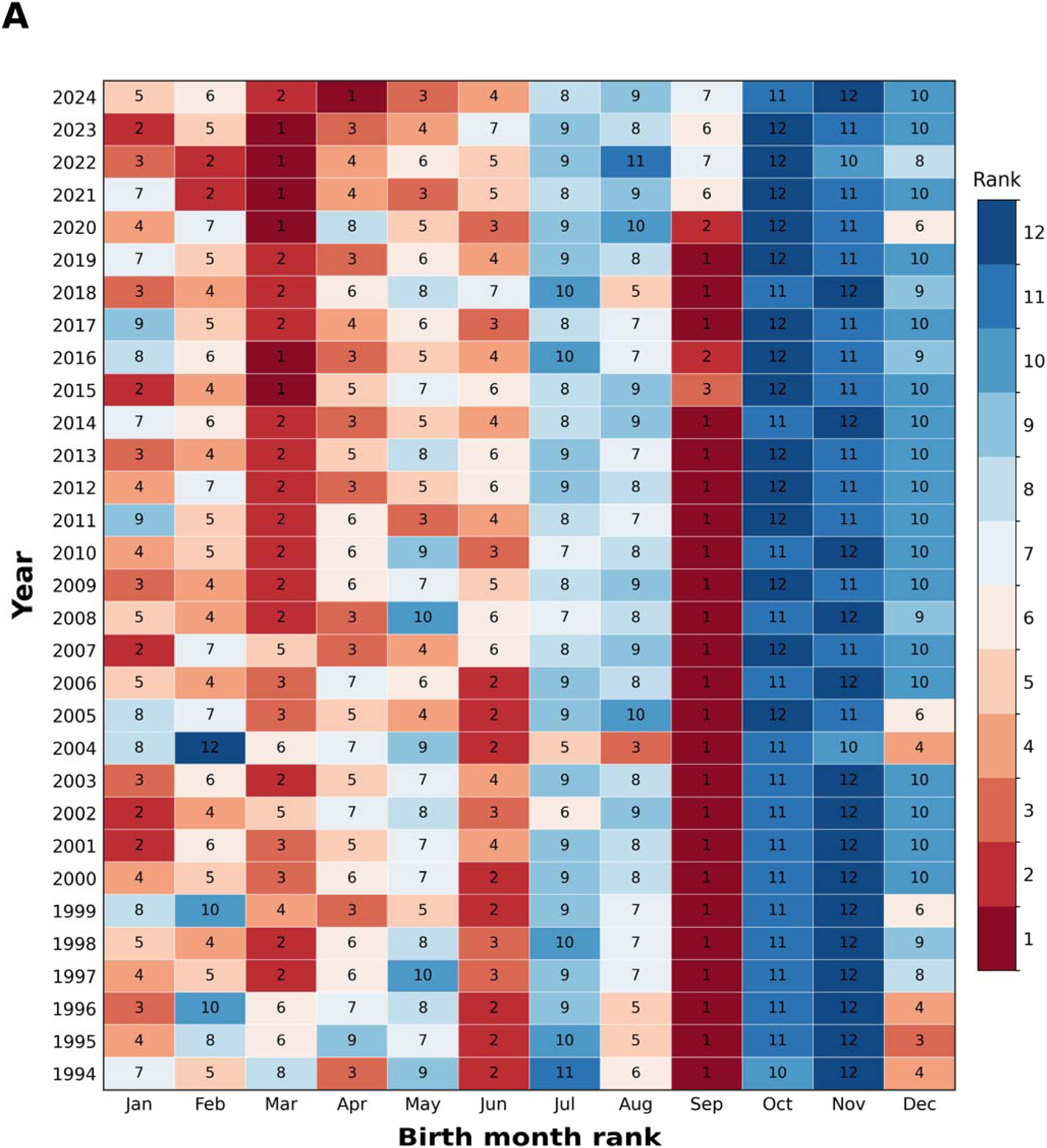
Birth month rank heatmap, South Africa, 1994-2024. Months are ranked within each year according to average daily recorded live births. Rank 1 indicates the month with the highest birth intensity. The figure shows the historical dominance of September births and the later emergence of March and April peaks.

**Figure 1B.**
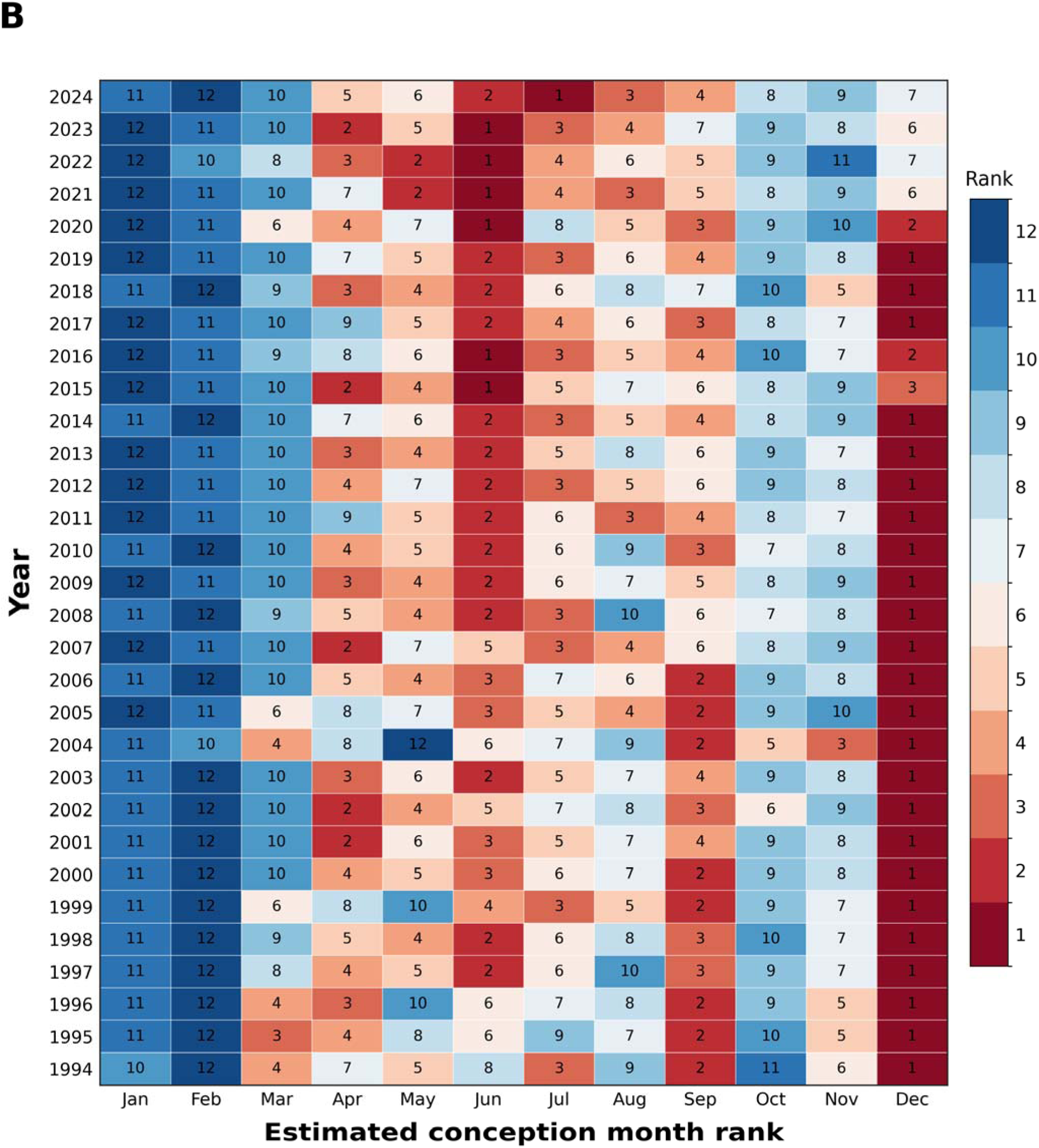
Estimated conception month rank heatmap, South Africa, 1994-2024. Estimated conception month ranks were obtained by shifting birth months approximately nine months earlier. The figure shows the transition from a December-dominant conception pattern to increasing June–July conception prominence.

The pattern began to weaken after 2014. March became the highest-ranked birth month in 2015 and 2016, although September again ranked highest in 2017, 2018 and 2019. March was again the highest-ranked month in 2020. From 2021 onwards, September no longer ranked first in any year; March was the highest-ranked month from 2021 to 2023, and April was highest in 2024.

Month-specific rank comparisons supported this descriptive reorganisation. Compared with 1994–2020, March became significantly more prominent from 2021 onwards (Wilcoxon rank-sum test, p = 0.013), while September became significantly less prominent (p < 0.001). April also became more prominent, although this did not reach conventional statistical significance (p = 0.054). When March, April and September were analysed together, the combined rank distribution did not differ significantly between periods (p = 0.518), reflecting changes in opposite directions across the individual months.

### Sex ratio at birth

The pre-pandemic monthly SRB series (January 2015 to December 2019) was stationary according to the Augmented Dickey-Fuller test (ADF statistic = −7.046, p < 0.001). Among the candidate autoregressive moving average (ARMA) models with autoregressive and moving-average orders ranging from 0 to 5, the ARMA(4,3) model had the lowest Akaike Information Criterion (AIC = −40.80) and was therefore selected (Supplementary Table 1).

Forecasts generated from the selected ARMA model identified four months between January 2021 and December 2021 with observed SRBs outside the 95% prediction intervals (Table 1; Fig. 2). The principal sustained deviation occurred during May through July 2021, when all three consecutive months fell below the lower 95% prediction bound.

**Figure 2.**
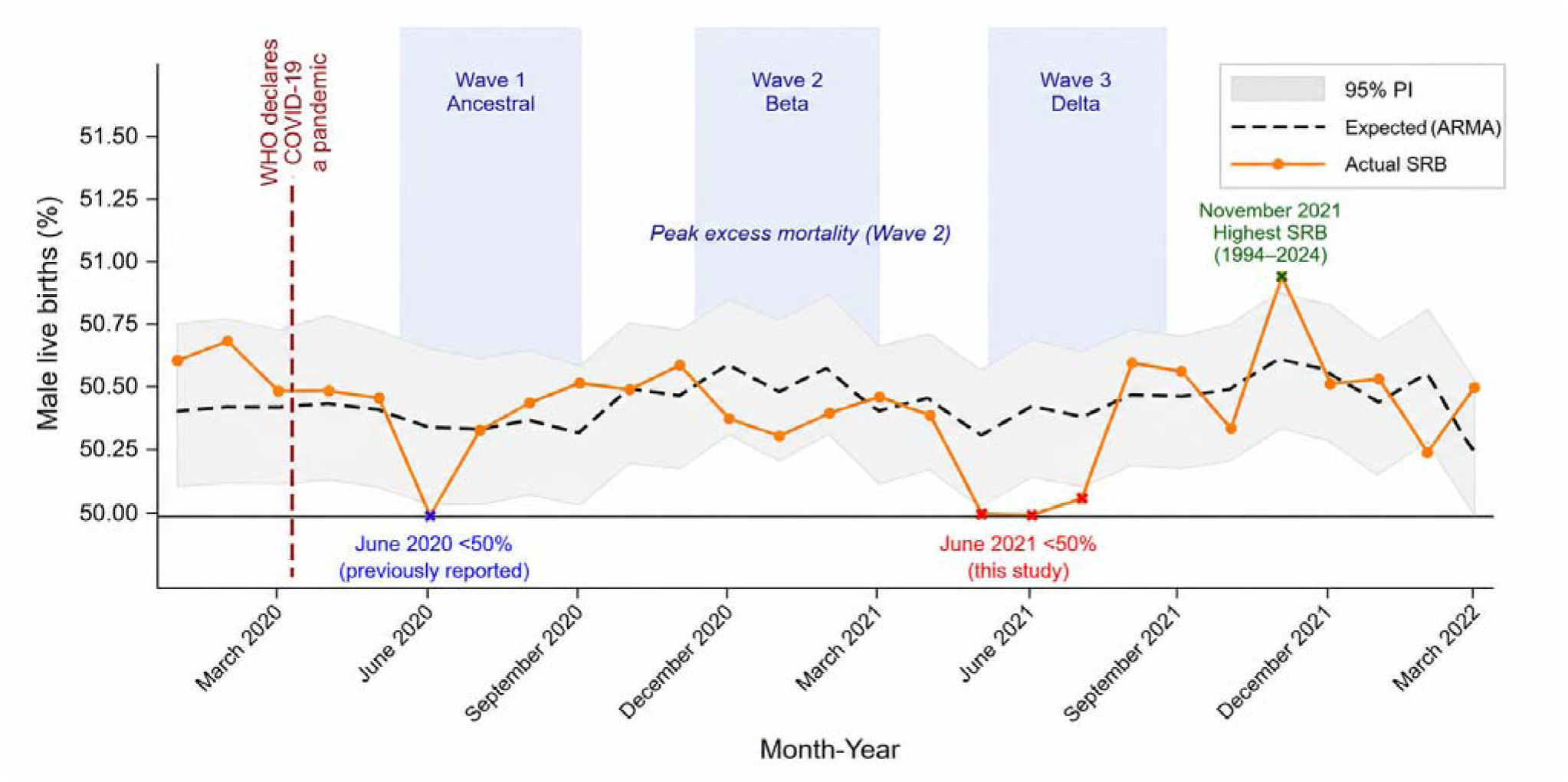
Sex ratio at birth from January 2020 to March 2022. Observed monthly male live births as a percentage of total live births (SRB) are shown with (autoregressive moving average) ARMA expected values and 95% prediction intervals (PI). The June 2020 inversion and May-July 2021 depression are marked. The complete January 2020–December 2024 forecast is shown in Supplementary Fig. 1.

**Table 1.**
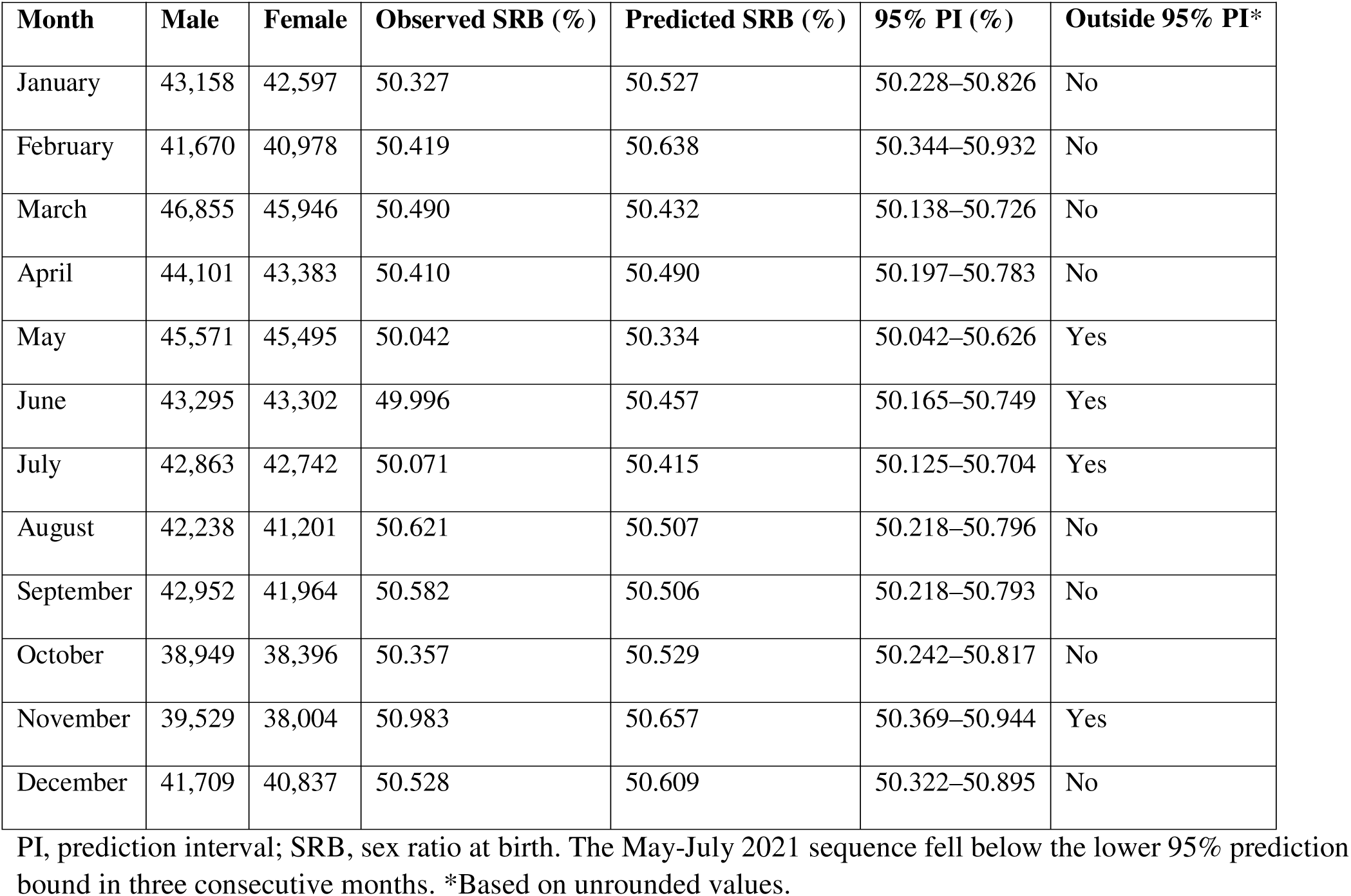
Observed and predicted sex ratio at birth for each month of 2021.

| Month | Male | Female | Observed SRB (%) | Predicted SRB (%) | 95% PI (%) | Outside 95% PI* |
| --- | --- | --- | --- | --- | --- | --- |
| January | 43,158 | 42,597 | 50.327 | 50.527 | 50.228–50.826 | No |
| February | 41,670 | 40,978 | 50.419 | 50.638 | 50.344–50.932 | No |
| March | 46,855 | 45,946 | 50.490 | 50.432 | 50.138–50.726 | No |
| April | 44,101 | 43,383 | 50.410 | 50.490 | 50.197–50.783 | No |
| May | 45,571 | 45,495 | 50.042 | 50.334 | 50.042–50.626 | Yes |
| June | 43,295 | 43,302 | 49.996 | 50.457 | 50.165–50.749 | Yes |
| July | 42,863 | 42,742 | 50.071 | 50.415 | 50.125–50.704 | Yes |
| August | 42,238 | 41,201 | 50.621 | 50.507 | 50.218–50.796 | No |
| September | 42,952 | 41,964 | 50.582 | 50.506 | 50.218–50.793 | No |
| October | 38,949 | 38,396 | 50.357 | 50.529 | 50.242–50.817 | No |
| November | 39,529 | 38,004 | 50.983 | 50.657 | 50.369–50.944 | Yes |
| December | 41,709 | 40,837 | 50.528 | 50.609 | 50.322–50.895 | No |

PI, prediction interval; SRB, sex ratio at birth. The May-July 2021 sequence fell below the lower 95% prediction bound in three consecutive months. \*Based on unrounded values.

Observed SRBs were 50.042% in May 2021, 49.996% in June 2021 and 50.071% in July 2021, compared with expected values of 50.334%, 50.457% and 50.415%, respectively (Table 1). The corresponding 95% prediction intervals were 50.042–50.626%, 50.165–50.749% and 50.125–50.704%. June 2021 was one of only two months between January 2020 and December 2024 in which the SRB fell below 50%; the other was June 2020. In a national population of this size, such inversion is uncommon, and all three months lay below the lower 95% prediction interval. Combining the monthly probabilities using Fisher’s method yielded χ² = 26.31 (6 degrees of freedom), p < 0.001. The observed SRB in November 2021 was 50.983%. The corresponding 95% prediction interval was 50.369–50.944% (Table 1). This represented the highest monthly SRB recorded during the entire 1994–2024 study period.

### Annual recorded live births

Annual recorded live births increased from 988,069 in 1994 to a peak of 1,112,378 in 2008, before entering a prolonged decline. The historical linear trend model estimated that annual births would have remained relatively stable, increasing modestly from 1,025,172 in 2020 to 1,030,356 in 2024. Observed births closely matched expectations in 2020 (1,021,561) and 2021 (1,017,735), but subsequently declined to 953,899 in 2022, 872,765 in 2023 and 798,556 in 2024. Annual births in 2022–2024 fell below the lower bound of the model’s 95% confidence interval, indicating that the post-2021 decline was large relative to this historical benchmark (Table 2; Fig. 3).

**Figure 3.**
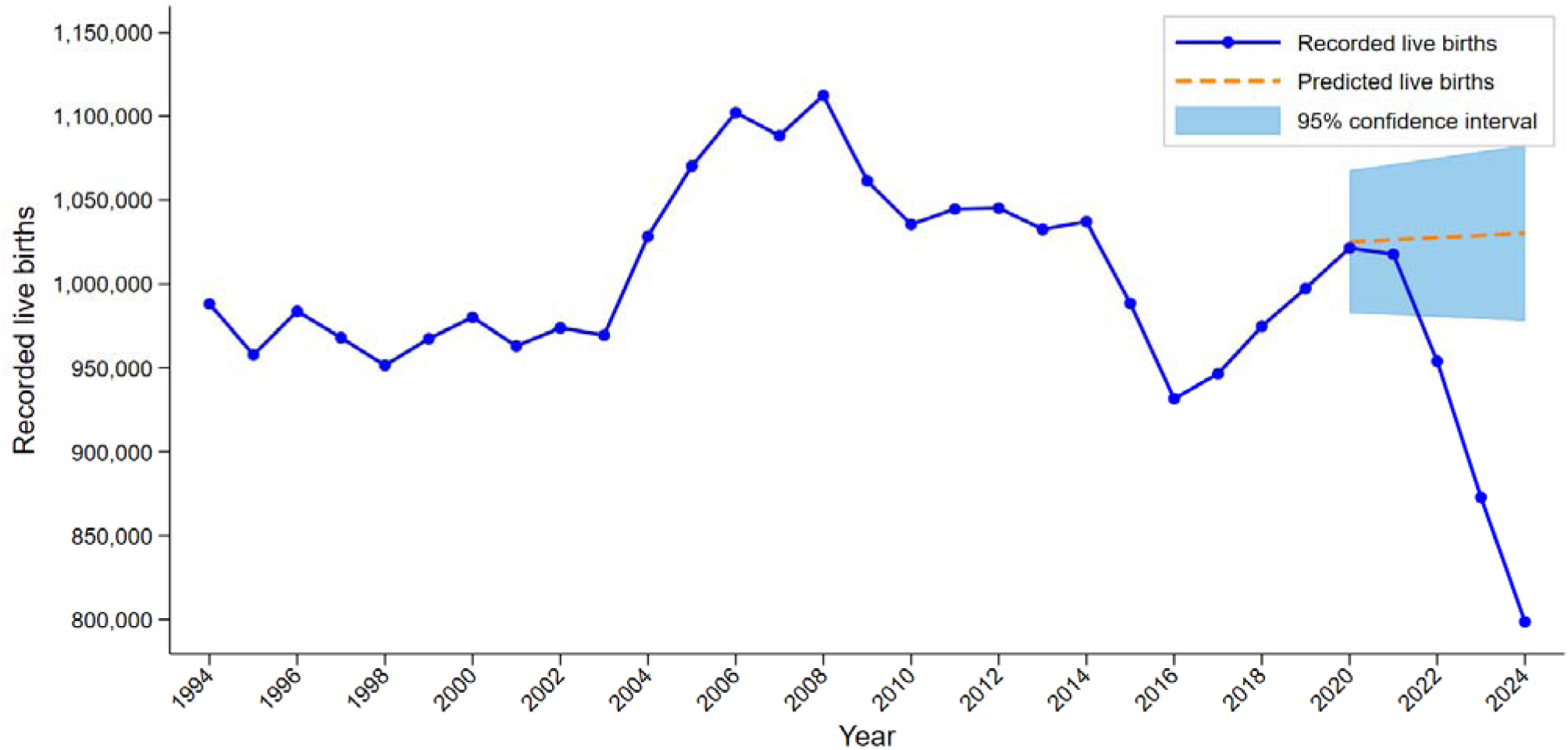
Annual recorded live births, expected live births and 95% confidence interval, South Africa, 1994-2024. Births after 2021 diverged markedly from expected trajectories, with the lowest observed value in 2024.

**Table 2.** Annual recorded live-birth landmarks, South Africa, 1994-2024.

| Year | Recorded live births | Change from 2021 | Expected births (95% CI) |
| --- | --- | --- | --- |
| 1994 | 988,069 | -2.9% | - |
| 2008 | 1,112,378 | 9.3% | - |
| 2020 | 1,021,561 | 0.4% | 1,025,172 (982,643-1,067,700) |
| 2021 | 1,017,735 | Reference | 1,026,468 (981,512-1,071,424) |
| 2022 | 953,899 | -6.3% | 1,027,764 (980,345-1,075,183) |
| 2023 | 872,765 | -14.2% | 1,029,060 (979,148-1,078,972) |
| 2024 | 798,556 | -21.5% | 1,030,356 (977,925-1,082,788) |

Expected annual births are model estimates; 95% confidence intervals (CI) are shown in parentheses.

## Discussion

This study identified three major demographic signals in South Africa between 1994 and 2024. First, the long-standing September birth peak, consistent with conceptions during the December–January holiday period, gave way after 2015 to increasingly prominent March and April birth peaks, indicating a shift towards winter conceptions. Second, the SRB exhibited a sustained decline between May and July 2021, including an inversion below 50% in June 2021. Third, annual recorded live births declined from a peak in 2008 to their lowest observed level in 2024, with the rate of decline accelerating after 2021. Although these findings collectively indicate substantial changes in South Africa’s reproductive demography over the past three decades, each represents a distinct demographic signal with potentially different underlying determinants and should therefore be interpreted independently rather than as components of a single causal pathway.

### From Christmas–New Year conceptions to winter conceptions

The birth seasonality findings are striking because of their historical continuity and subsequent change. The September peak reported by Cowgill and Lam and Miron persisted into the democratic era (Cowgill, 1966, Lam and Miron, 1991). The first two decades of the series therefore support the interpretation that December and early January conceptions remained a stable feature of South African reproductive timing. Similar holiday-related birth patterns have been described elsewhere. In France, a concentration of births around late September has been interpreted as reflecting New Year conceptions (Régnier-Loilier, 2010).

The later emergence of March and April peaks suggests a meaningful shift in reproductive timing. March births correspond approximately to June conceptions, while April births correspond approximately to July conceptions. The mechanism cannot be inferred directly from birth registration data. Plausible contributors include changing household electrification, indoor comfort, work and leisure rhythms, urbanisation, social norms, contraception and environmental conditions. Within South Africa, increasing urbanisation and structural changes in labour markets are likely to have contributed to a gradual weakening of traditional seasonal fertility patterns, consistent with a shift toward more individually planned reproductive timing (Dorélien, 2016).

Temperature may also matter, as prior work has linked heat shocks to dynamic changes in birth rates, effectively redistributing births away from hotter periods (Barreca et al., 2018). However, because temperature was not modelled directly, whether rising temperatures associated with climate change contributed to the observed shift from summer (December) to winter (June-July) conceptions remains speculative. More generally, environmental influences on reproductive timing are well documented but context-dependent, and the present data cannot disentangle specific drivers.

This caveat is reinforced by recent evidence that high temperatures in the months before birth are associated with fewer male births in sub-Saharan Africa and India (Abdel Ghany et al., 2026), and by historical evidence from Poznań that ambient temperature was associated with variation in the SRB (Liczbińska et al., 2024). Although these studies concern offspring sex ratios rather than conception timing directly, they support the broader premise that ambient temperature can influence human reproductive patterns and therefore provide relevant context for the observed shift from December holiday conceptions towards June–July winter conceptions. The shift also resonates with familiar South African popular references to winter intimacy, including public references to winter cuddles (Duchess of Healing, 2019), which should be interpreted as culturally descriptive narratives rather than evidence of behavioural causation.

### The May–July 2021 sex ratio at birth perturbation

The May–July 2021 SRB deviation is the most acute signal in the analysis. It occurred three to five months after the January 2021 mortality peak associated with the Beta wave, which is temporally consistent with the stress-sensitive window described in the sex-ratio literature (Masukume and Grech, 2015, Masukume et al., 2022). This timing is important. It links the observed births not to the onset of the pandemic, which was followed by the June 2020 inversion already reported (Masukume et al., 2022), but to the later and more severe Beta wave. The fact that the June 2021 SRB fell below 50% is also important. Chao and colleagues showed that SRB is usually above parity, with country and regional reference levels estimated around the familiar male excess at birth (Chao et al., 2019). Therefore, a monthly SRB below 50% in a national population of South Africa’s size should not be dismissed as a trivial fluctuation merely because it is close to parity; it is already notably low before formal statistical testing is considered. A large body of literature demonstrates that the SRB is sensitive to acute population-level stressors, with consistent transient declines reported following epidemics, natural disasters, economic crises, and other major societal shocks, typically with a lag of approximately 3–5 months (Masukume and Grech, 2026). However, the magnitude and detectability of these effects vary across contexts, and null or weak findings are also common, particularly in large, heterogeneous populations (Masukume and Grech, 2026).

The persistence of the South African signal into 2021 is notable in an international context. In England and Wales, COVID-19 was associated with a June 2020 decline in the SRB and a subsequent December 2020 increase, but not a comparable sustained perturbation during 2021 (Masukume et al., 2023). In the Republic of Ireland and Northern Ireland, COVID-19-related SRB disruptions were also centred on 2020 rather than 2021 (Masukume et al., 2024, Masukume et al., 2025). By contrast, analyses from the United States and Finland did not detect significant pandemic-related changes in SRB (Cleaver and Non, 2024, Helle et al., 2026). The South African May-July 2021 depression therefore appears unusual both because of its duration and because it occurred after the pandemic’s onset, coinciding instead with the exceptionally severe Beta-wave mortality period.

Several independent sources support the interpretation that early 2021 was a period of severe population stress in South Africa and the wider southern African region. National excess-mortality analyses showed that the second wave produced the greatest mortality burden of the first four waves in South Africa (Bradshaw et al., 2022). The Beta variant was first described in South Africa and became the dominant lineage during the second wave (Tegally et al., 2021). Hospital-based analyses found higher mortality in the second wave than in the first (Jassat et al., 2021), and later work described the Beta variant as particularly virulent in relation to mortality and case fatality (Hirachund et al., 2024). Evidence from mental health and reproductive health also triangulates the timing: nationally representative panel data recorded elevated depressive symptoms in February 2021 (Dladla-Jaca et al., 2024), and a national review of maternal, perinatal and reproductive health reported marked disruptions during the COVID-19 period, including a peak in stillbirth burden around the January 2021 Beta-wave mortality surge (Fawcus et al., 2024). The present findings extend this literature by suggesting that in South Africa the strongest SRB perturbation may not coincide with the initial pandemic shock (2020), but rather with the subsequent Beta-wave mortality peak in early 2021. This interpretation remains associative, as the available data do not permit direct identification of causal mechanisms or individual-level exposure pathways. The SRB perturbation should therefore be interpreted as a short-term stress-sensitive demographic signal.

Scholarly obituaries and tributes documenting the deaths of prominent clinicians and medical researchers in the region during the Beta-wave period further illustrate the exceptional salience of mortality among health professionals and public figures at that time, reinforcing the plausibility that the Beta wave was experienced as a profound societal stressor beyond ordinary epidemic statistics (Green, 2021a, Green, 2021b, Ray, 2021, Watts, 2021). These sources provide contextual corroboration of societal stress intensity, although they do not constitute direct evidence of demographic mechanism.

### The November 2021 increase in the sex ratio at birth

November 2021 represented the highest monthly SRB observed during the 31-year study period and exceeded the upper 95% prediction interval. This occurred approximately nine months after the national transition that followed the January 2021 Beta-wave mortality peak. On 1 February 2021, South Africa’s President announced that the first COVID-19 vaccine shipment had arrived, that average daily infections had almost halved, that hospital admissions were falling, and that the country had passed the peak of the second wave (South African Government, 2021). The same address announced easing under adjusted Alert Level 3, including reopening beaches, parks and public swimming pools, extending curfew hours, permitting faith-based gatherings, and easing alcohol sales restrictions (Mapanga et al., 2023). The speech explicitly framed this moment as a new chapter, stating that the vaccine arrival contained “the promise that we can turn the tide” and ending with a call to “keep the flame of hope alive”.

This timing raises the hypothesis that renewed optimism and increased opportunities for social interaction after a period of acute restriction and mortality stress may have increased partnered sexual activity and consequently conceptions during February 2021.

Such a behavioural pathway is biologically plausible, as Guerrero demonstrated a U-shaped relationship between coital frequency and the SRB, whereby higher coital frequency was associated with an increased probability of male conception (Guerrero, 1974). However, the SRB should not be interpreted as a direct proxy for sexual activity. Similar SRB increases approximately nine months after the March 2020 pandemic declaration were reported in England and Wales and Northern Ireland, where they were interpreted as potentially reflecting increased partnered sexual activity in a subset of the population during lockdown onset; however, England and Wales and Northern Ireland also recorded the lowest monthly births in December 2020 (Masukume et al., 2023, Masukume et al., 2025). South Africa may therefore represent a different pattern: a post-Beta-wave relaxation signal rather than a lockdown-onset signal. This distinction is important because the November 2021 increase followed a period characterised by declining mortality, vaccine availability, and gradual easing of social restrictions, rather than the initial disruption associated with pandemic emergence. This interpretation remains hypothesis-generating and requires further evaluation using individual-level reproductive behaviour data, mobility indicators, and fertility trends. The present discussion focuses on the most prominent and biologically notable SRB signals identified in the study.

### Fertility decline 2022–2024

The annual live-birth decline is a separate process. Births peaked in 2008 and then generally declined, indicating that fertility change was already underway before COVID-19. This long-term decline is consistent with established demographic transition theory and sustained fertility reductions observed in middle-income countries undergoing urbanisation, educational expansion, and labour market restructuring (Bongaarts, 2002, Bryant, 2007). The acceleration after 2021, however, is notable. The decline from 2021 to 2024 is consistent with a possible delayed fertility response to the severe mortality and uncertainty of the 2021 waves rather than an immediate response to the initial 2020 shock. Similar delayed fertility responses following major societal disruptions have been documented elsewhere and are often interpreted in terms of postponement effects alongside potential medium-term quantum adjustments (Goldstein et al., 2013, Sobotka et al., 2011).

This pattern differs from many high-income countries, where fertility responses often appeared earlier, with birth troughs in early 2021 followed by partial rebounds (Pomar et al., 2022, Sobotka et al., 2024). South Africa appears to have experienced a delayed and sustained decline. This pattern is broadly consistent with cross-country evidence showing that fertility responses to the COVID-19 pandemic were heterogeneous and shaped by differences in economic vulnerability, labour market insecurity, and access to social protection (Comolli and Vignoli, 2021, Sobotka et al., 2024). Economic uncertainty, partnership disruption, health-service disruption, migration, caregiving burdens and changing fertility preferences may all have contributed. Recent work on the later phase of the pandemic emphasises the roles of policy interventions, vaccination programmes and economic uncertainty in shaping fertility responses (Winkler-Dworak et al., 2024). In lower- and middle-income settings, weaker social protection systems and more precarious labour market conditions may amplify such effects, potentially contributing to more persistent fertility declines rather than short-term postponement followed by full recovery. The continued decline through 2023 and 2024 suggests that the reduction was not simply a short-term postponement followed by recovery. Some births may have been foregone rather than merely delayed, although this distinction cannot be determined from aggregate live-birth data alone.

### Comparative implications

An important question is whether similar demographic perturbations occurred elsewhere in southern Africa during the Beta-driven second wave of COVID-19. Although first identified in South Africa (Tegally et al., 2021), the Beta variant also predominated in neighbouring Zimbabwe, accounting for approximately 95% of sequenced SARS-CoV-2 samples during the second-wave peak, which was associated with higher adjusted in-hospital mortality than the first wave (Mashe et al., 2021, Fryatt et al., 2024). Beta transmission was likewise documented in Zambia and Mozambique (Mwenda et al., 2021, Martínez-Martínez et al., 2023). Comparative analyses of monthly live births and the SRB across southern African countries would help determine whether the perturbation observed in South Africa between May and July 2021 reflects a country-specific phenomenon or a broader regional demographic response to the Beta wave.

The South African findings raise a comparative question for other large middle-income countries that experienced severe pandemic waves. Brazil, like South Africa, is classified as an upper-middle-income economy, whereas India is classified as lower-middle-income; all three therefore sit within the broad middle-income group rather than among high-income countries (World Bank Group, 2025). Brazil experienced a major Gamma-associated epidemic wave, with genomic and epidemiological work showing that the P.1/Gamma lineage emerged in Manaus and was associated with a large resurgence despite high prior attack rates (Faria et al., 2021). India experienced a devastating second wave in 2021, and genomic epidemiology identified Delta/B.1.617.2 as a major driver of the Delhi surge (Dhar et al., 2021). Excess mortality estimates suggest that this wave represented one of the more severe pandemic mortality events globally, although with substantial uncertainty across models but consistently high estimated burdens (Anand et al., 2021, Wang et al., 2022). Notably, increased stillbirths were also documented during the Gamma- and Delta-dominated waves in Brazil and India, respectively, supporting investigation of broader reproductive-demographic perturbations beyond South Africa (Dandona et al., 2023, Xavier et al., 2023).

These countries are not direct analogues of South Africa, but their large populations, middle-income health-system contexts and severe variant-driven waves make them important comparators. From a demographic perspective, both Brazil and India experienced notable fertility disruptions during the pandemic period, although the timing and direction of change varied across regions, consistent with heterogeneous fertility responses observed globally (Comolli and Vignoli, 2021, Sobotka et al., 2024). However, relatively limited research has examined SRB dynamics in these settings during COVID-19, particularly at monthly resolution. Existing SRB studies remain concentrated in high-income countries, where effects have generally been small, transient, or absent (Helle et al., 2026). This highlights a potential empirical gap for large middle-income populations that experienced substantial pandemic-related mortality shocks. A future analysis of monthly SRB and live-birth trajectories in Brazil, India and other countries could help determine whether the delayed South African 2021 SRB depression was a country-specific signal or part of a broader pattern in populations exposed to intense pandemic stress. Such comparative work would also allow investigation of whether SRB perturbations may align more closely with peak mortality timing (e.g., Gamma in Brazil, Delta in India) rather than with initial pandemic onset, as tentatively suggested by the South African findings. More broadly, cross-country demographic research on COVID-19 has shown that the magnitude of fertility and mortality responses is strongly conditioned by institutional capacity, inequality, and the severity and timing of epidemic waves, suggesting that middle-income countries may exhibit qualitatively different temporal patterns compared with high-income settings (Aburto et al., 2021, Sobotka et al., 2024).

### Practical implications of changing birth volumes

Changes in the absolute number of births have practical implications beyond demographic description. Maternity-unit staffing models necessarily depend on delivery volume, acuity and the number and mix of staff physically present; a recent scoping review proposed a core delivery-unit staffing indicator calculated from annual births divided by days and by average shift staffing (Stones and Nair, 2023). Declining or shifting birth volumes therefore may affect the planning of labour wards, postnatal beds, neonatal services, theatre capacity and community midwifery. In health systems with constrained capacity, even moderate changes in annual birth counts can have implications for service organisation, particularly where staffing levels cannot be rapidly adjusted. Birth numbers also matter for immunisation planning because vaccine coverage calculations and dose forecasting depend on the size of the infant target population; WHO and UNICEF methods use live births or surviving infants as key denominators for infant vaccination coverage (Burton et al., 2009). The sharp post-2021 decline in recorded births may therefore have potential consequences for resource allocation, workforce planning and procurement, while also requiring caution so that apparent reductions in workload do not obscure changes in case complexity or inequitable access to care. In particular, aggregate declines in live births may mask increased heterogeneity in maternal risk profiles, which can increase per-birth resource intensity even when total volume falls. More broadly, abrupt changes in fertility and birth volume can create planning lag effects, as health systems typically adjust staffing and infrastructure based on multi-year trends rather than short-term demographic shocks.

### Strengths and limitations

The main strength of this study is its use of national recorded live-birth data covering 31 years and 372 monthly observations. Standardising by days per month avoids misleading interpretation of raw monthly counts. The rank-based seasonality analysis provides a simple and robust way to show changes in the full monthly structure. The SRB analysis uses a recent pre-pandemic baseline and explicit prediction intervals. A second strength is the integrated interpretation of seasonality, SRB and annual births while keeping their mechanisms distinct.

The study also has limitations. It is ecological and cannot establish individual-level mechanisms. Gestational timing is approximated at population level. Registration completeness and delays may affect counts, although the stability and magnitude of the observed patterns make it unlikely that the findings are artefacts of reporting or data processing. The study did not directly model temperature, electricity access, age-specific fertility, parity, province, migration, contraception or socioeconomic conditions. As a result, observed associations should be interpreted as descriptive rather than mechanistic, and potential confounding pathways remain unquantified. Causal language has therefore been avoided. The 2024 recorded live-birth total may be particularly sensitive to release timing and registration lags, and should be rechecked when later Statistics South Africa releases become available.

## Conclusion

Between 1994 and 2024, South Africa experienced major changes in reproductive timing, sex-ratio patterning and annual recorded live births. The long-standing September birth peak, consistent with Christmas holiday and New Year conceptions, gave way after 2015 to increasing March and April peaks, consistent with more winter conceptions. The COVID-19 period included a rare May–July 2021 depression in the SRB, including a June 2021 inversion below 50%, temporally aligned with the aftermath of the January 2021 Beta-wave mortality peak and supported by independent evidence of societal, mental-health and perinatal stress. In contrast, November 2021 recorded the highest monthly SRB observed during the 31-year study period, approximately nine months after the easing of COVID-19 restrictions in February 2021. Annual recorded live births declined from their 2008 peak to the lowest observed level in 2024, with acceleration after 2021.

Although this ecological study cannot establish causal pathways linking these processes, the findings show that birth seasonality, SRB and annual live births can be interpreted together as complementary indicators of population reproduction while preserving their distinct biological and demographic mechanisms. Collectively, these findings demonstrate that changes in population reproductive patterns provide important insights into how populations respond to major societal and public-health change.

## Data Availability

All data produced are available online at https://www.statssa.gov.za/?page_id=1854&PPN=P0305

### Data availability

The recorded live-birth data used in this study are publicly available from Statistics South Africa recorded live birth releases and appendices https://www.statssa.gov.za/?page_id=1854&PPN=P0305.

### Ethics statement

This study used anonymised aggregate public-domain data. No individual-level data were analysed. Ethics approval was not required.

### Funding

No specific funding was received for this study.

### Author contributions

Conceptualisation: RM, PTC, GM, GL, VG and WM. Data curation: GM. Formal analysis: RM, GM and VG. Methodology: RM and GM. Visualisation: RM and GM. Writing - original draft: GM and GL. Writing - review and editing: RM, PTC, GM, GL, VG and WM. All authors approved the final manuscript.

### AI Use Disclosure

ChatGPT (OpenAI) was used for language editing.

