## Supplementary appendix for "From Christmas sex to winter intimacy: three decades of birth seasonality, sex ratio dynamics, and fertility change in South Africa, 1994–2024"

**Supplementary Table 1**

Parameter selection procedure for the autoregressive moving average (ARMA) model to fit the sex ratio at birth in South Africa.

| **AR** | **MA** | **AR L1** | **AR L2** | **AR L3** | **AR L4** | **AR L5** | **MA L1** | **MA L2** | **MA L3** | **MA L4** | **MA L5** | **AIC** |
| --- | --- | --- | --- | --- | --- | --- | --- | --- | --- | --- | --- | --- |
| 0 | 0 |  |  |  |  |  |  |  |  |  |  | -33.22 |
| 0 | 1 |  |  |  |  |  | 0.06 |  |  |  |  | -31.48 |
| 0 | 2 |  |  |  |  |  | 0.01 | 0.20 |  |  |  | -30.35 |
| 0 | 3 |  |  |  |  |  | 0.32 | 0.03 | 0.56 |  |  | -33.08 |
| 0 | 4 |  |  |  |  |  | 0.12 | -0.16 | 0.16 | -0.55 |  | -39.94 |
| 0 | 5 |  |  |  |  |  | 0.08 | -0.34 | 0.11 | -0.69 | -0.16 | -39.01 |
| 1 | 0 | 0.07 |  |  |  |  |  |  |  |  |  | -31.51 |
| 1 | 1 | NC | NC | NC | NC | NC | NC | NC | NC | NC | NC | NC |
| 1 | 2 | 0.79 |  |  |  |  | -0.84 | -0.16 |  |  |  | -31.42 |
| 1 | 3 | -0.52 |  |  |  |  | 0.77 | 0.21 | 0.44 |  |  | -36.59 |
| 1 | 4 | 0.36 |  |  |  |  | -0.29 | -0.28 | 0.23 | -0.66 |  | -40.21 |
| 1 | 5 | 0.62 |  |  |  |  | -0.54 | -0.26 | 0.30 | -0.70 | 0.20 | -38.93 |
| 2 | 0 | 0.06 | 0.07 |  |  |  |  |  |  |  |  | -29.84 |
| 2 | 1 | 0.98 | -0.19 |  |  |  | -1.00 |  |  |  |  | -32.02 |
| 2 | 2 | 0.09 | 0.59 |  |  |  | 0.00 | -1.00 |  |  |  | -34.84 |
| 2 | 3 | 0.06 | 0.59 |  |  |  | 0.04 | -1.00 | -0.04 |  |  | -32.90 |
| 2 | 4 | 0.24 | 0.24 |  |  |  | -0.17 | -0.43 | 0.17 | -0.57 |  | -39.72 |
| 2 | 5 | 0.09 | 0.30 |  |  |  | -0.02 | -0.48 | 0.13 | -0.52 | -0.12 | -37.82 |
| 3 | 0 | 0.06 | 0.07 | 0.08 |  |  |  |  |  |  |  | -28.17 |
| 3 | 1 | 0.93 | 0.02 | -0.23 |  |  | -1.00 |  |  |  |  | -32.92 |
| 3 | 2 | 0.11 | 0.59 | -0.05 |  |  | -0.00 | -1.00 |  |  |  | -32.99 |
| 3 | 3 | -0.88 | 0.65 | 0.66 |  |  | 0.98 | -0.98 | -1.00 |  |  | -33.20 |
| 3 | 4 | 0.74 | 0.19 | -0.39 |  |  | -0.71 | -0.43 | 0.71 | -0.57 |  | -39.53 |
| 3 | 5 | 1.13 | 0.00 | -0.57 |  |  | -1.13 | -0.18 | 1.04 | -0.60 | 0.31 | -39.15 |
| 4 | 0 | 0.08 | 0.11 | 0.10 | -0.41 |  |  |  |  |  |  | -36.44 |
| 4 | 1 | 0.62 | 0.06 | 0.07 | -0.42 |  | -0.68 |  |  |  |  | -37.63 |
| 4 | 2 | 0.10 | 0.76 | -0.02 | -0.41 |  | -0.03 | -0.97 |  |  |  | -40.39 |
| 4 | 3 | 0.68 | 0.48 | -0.51 | -0.30 |  | -0.74 | -0.74 | 1.00 |  |  | -40.80 |
| 4 | 4 | 0.40 | 0.74 | -0.40 | -0.53 |  | -0.43 | -0.99 | 0.76 | 0.32 |  | -39.51 |
| 4 | 5 | 0.70 | 0.39 | -0.39 | -0.36 |  | -0.71 | -0.56 | 0.79 | -0.13 | 0.23 | -37.99 |
| 5 | 0 | 0.03 | 0.12 | 0.12 | -0.40 | -0.13 |  |  |  |  |  | -35.27 |
| 5 | 1 | 0.70 | 0.06 | 0.06 | -0.45 | 0.08 | -0.75 |  |  |  |  | -35.78 |
| 5 | 2 | -0.15 | 0.65 | 0.11 | -0.43 | -0.23 | 0.24 | -0.76 |  |  |  | -39.11 |
| 5 | 3 | 0.73 | 0.54 | -0.58 | -0.39 | 0.14 | -0.75 | -0.75 | 1.00 |  |  | -39.69 |
| 5 | 4 | 1.27 | 0.32 | -0.87 | -0.20 | 0.38 | -1.30 | -0.55 | 1.36 | -0.40 |  | -38.28 |
| 5 | 5 | 0.58 | 0.36 | -0.25 | -0.37 | -0.12 | -0.60 | -0.53 | 0.62 | -0.10 | 0.34 | -36.06 |

AIC: Akaike Information Criterion; AR: autoregressive parameter; MA: moving average parameter; NC: not converged. The minimum AIC was obtained for ARMA(4,3) (AIC = -40.80).

**Supplementary Figure 1**

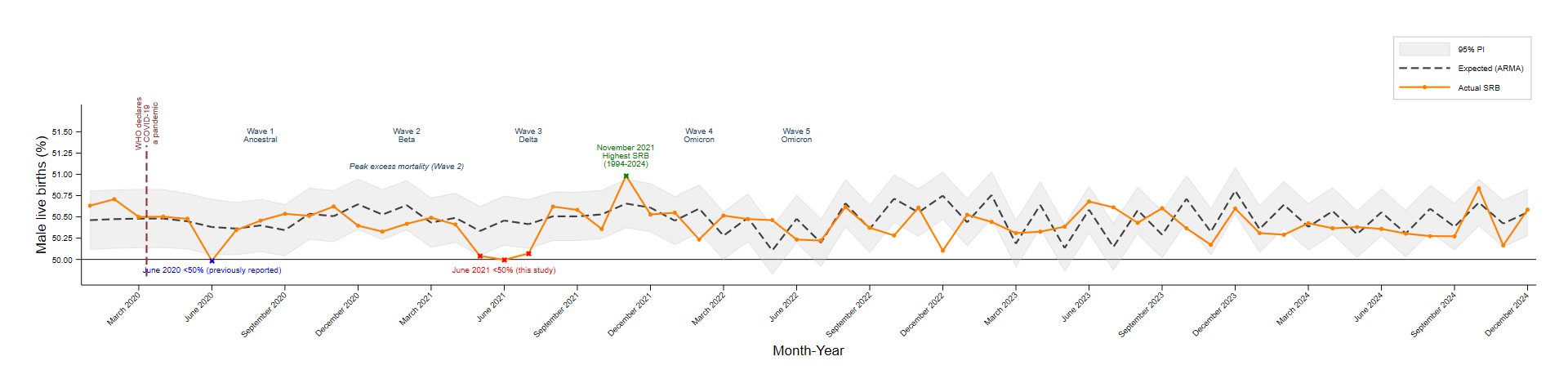

PI, prediction intervals; ARMA (autoregressive moving average).
